# Closed-loop stimulation of lateral cervical spinal cord in upper-limb amputees to enable sensory discrimination

**DOI:** 10.1101/2022.03.25.22272820

**Authors:** Ameya C. Nanivadekar, Santosh Chandrasekaran, Eric R. Helm, Michael L. Boninger, Jennifer L. Collinger, Robert A. Gaunt, Lee E. Fisher

**Affiliations:** Rehab Neural Engineering Labs, University of Pittsburgh, Pittsburgh, PA 15213; Department of Bioengineering, University of Pittsburgh, Pittsburgh, PA 15213; Department of Physical Medicine and Rehabilitation, University of Pittsburgh, Pittsburgh, PA 15213; Center for Neural Basis of Cognition, Pittsburgh, PA, 15213; Human Engineering Research Labs, VA Center of Excellence, Department of Veteran Affairs, Pittsburgh, PA, 15206; University of Pittsburgh Clinical Translational Science Institute, Pittsburgh, PA, 15213

**Author notes:** To whom correspondence should be addressed: Lee E. Fisher, 3520 Fifth Avenue, Suite 300, Pittsburgh, PA 15213. Authors ACN and SC contributed equally to this work.

## Abstract

Modern myoelectric prosthetic hands have multiple independently controllable degrees of freedom, but require constant visual attention to use effectively. As we know from motor control of our native limbs, somatosensory feedback is essential to control our movements and provides information not available through vision alone. Similarly, stimulation of the nervous system can potentially provide artificial somatosensory feedback to reduce the reliance on visual cues to efficiently operate prosthetic devices. We have shown previously that epidural stimulation of the lateral cervical spinal cord can evoke tactile sensations perceived as emanating from the missing arm and hand in people with upper-limb amputation. In this study, two subjects with upper-limb amputation used this somatotopically-matched tactile feedback to discriminate object size and compliance while controlling a prosthetic hand. With less than 30 minutes of training each day, both subjects were able to use artificial somatosensory feedback to perform a subset of the discrimination tasks at a success level well above chance. Subject 1 was consistently more adept at determining object size (74% accuracy; chance: 33%) while Subject 2 achieved a higher accuracy level in determining object compliance (60% accuracy; chance 33%). In each subject, discrimination of the other object property was only slightly above or at chance level suggesting that the task design and stimulation encoding scheme are important determinants of which object property could be reliably identified. Our observations suggest that artificial somatosensory feedback provided via spinal cord stimulation can be readily used to infer information about the real-world with minimal training, but that task design is critical and that performance improvements may not generalize across tasks.

## Introduction

Limb loss has a profound impact on the ability of individuals to interact with their environment and perform activities of daily living. In addition to learning to use a prosthetic limb, people with upper-limb amputation have to devise ways of compensating for the lack of sensory feedback, even with state-of-the-art prosthetic devices. The current repertoire of prosthetics range from simple cosmetic hands to cable actuated hooks and dexterous robotic limbs. However, none of these devices provide tactile feedback, and as a result, intuitive control remains elusive. In fact, users often prefer simpler body-powered prosthetics because they can infer information about limb state from pressure exerted on the residual limb ^1^. Individuals with upper-limb amputation also rely on sustained visual attention to compensate for the lack of somatosensory feedback ^2^. This reliance on visual cues leads to sub-optimal motor control in various situations such as attempting to grasp an object that is out of the line of sight, or rapidly modulating grip force to prevent object slipping^3^. Highlighting these challenges, surveys of upper-limb prosthesis users indicate restoring somatosensory feedback as a top unmet need^4–7^.

Normal haptic perception requires an interplay between tactile and proprioceptive modalities of sensory information^8^. Broadly, tactile sensation conveys information about object contact forces, temperature, and surface features of an object^8^. Meanwhile, proprioception conveys information about the state and orientation of the hand and fingers which enables stereognosis (i.e. inference of object location, shape, and size)^8^. Several studies have demonstrated the effectiveness of artificial somatosensory feedback in conveying these multiple modalities of information during prosthesis use. When tactile information was delivered by electrical stimulation of peripheral nerves in the residual limb, study participants demonstrated improvements in manipulating objects^9^, controlling grip force^10–12^, and identifying object compliance^13–16^. Further, electrical stimulation designed to mimic mechanoreceptor firing patterns enabled amputees to discriminate naturalistic textures^17,18^. More recently, feedback via peripheral nerve stimulation has been incorporated into a take-home system to demonstrate that subjects could learn to use this artificial somatosensory feedback in tasks of daily living^19^.

While tactile sensations are routinely evoked with electrical stimulation, reliable proprioceptive sensations have remained elusive. As a result, proprioceptive information has been conveyed by remapping the intensity of an evoked tactile sensation to a signal such as grasp aperture or finger joint angle. With these techniques, participants could discriminate object size with a success rate better than chance^14–16^.

Stimulation of the lateral cervical spinal cord can evoke focal sensations in the missing fingers and hand, even in people with high level amputations, such as at the proximal arm or shoulder ^20^. In addition to conveying information about location, increased the amplitude of spinal cord stimulation (SCS) led to linearly-modulated increases in percept intensity^20^. The primary goal of this study was to determine if artificial somatosensory feedback provided by SCS could provide functionally relevant information during control of a prosthetic limb. Two subjects with upper-limb amputation interacted with objects of varying size and compliance using a sensorized DEKA^21,22^ hand or a virtual representation of that hand, rendered in MuJoCo^23,24^. Somatotopically-matched tactile sensory feedback was provided via lateral SCS by varying the stimulus amplitude in real-time. Subjects were asked to determine the size or compliance of the object based on this feedback. For each subject, the utility of feedback via SCS was inferred from the performance on the object discrimination task. Additionally, we characterized features of the closed-loop control system and task design that affected the utility of this feedback and performance on the discrimination task.

## Methods

### Study design

The aim of this study was to use SCS to provide real-time sensory feedback so that subjects could use a sensorized prosthetic hand to interact with objects and determine their size and compliance. To characterize the factors affecting the utility of sensory feedback, subjects performed an object discrimination task in two different control environments (real and virtual reality).

Two subjects with upper-limb amputations were recruited for this study. Subject 1 had a trans-humeral amputation of the right arm. Subject 2 had a trans-radial amputation of the right arm, and also had a right hemisphere stroke that resulted in extensive paN/xralysis of the contralateral limb. The time since amputation was greater than two years for Subject 1 and three years for Subject 2. All procedures and experiments were approved by the University of Pittsburgh and Army Research Labs Institutional Review Boards and subjects provided informed consent before participation.

### Electrode implantation

SCS leads were implanted through a minimally invasive, outpatient procedure performed under local anesthesia, described previously^20^. Briefly, three 16-contact SCS leads (Infinion, Boston Scientific) were percutaneously inserted into the epidural space on the lateral aspect of the C5– C8 spinal cord. Contacts were 3 mm long, with 1 mm inter-contact spacing. Lead placement was iteratively adjusted based on the subjects’ verbal report of the location of sensations evoked by intraoperative stimulation. The leads were maintained for fewer than 29 days. Subjects attended testing sessions 3–4 days per week during the implantation period. For each subject, the majority of implant time was used to characterize perceptual characteristics of the evoked sensations^20^.

The functional closed-loop experiments described here were performed during the last 5 days of the study.

### Neural stimulation

During testing sessions, stimulation was delivered using three 32-channel stimulators (Nano 2+Stim; Ripple, LLC). The maximum current output for these stimulators was 1.5 mA per channel. To achieve the higher current amplitudes required for SCS, a custom-built circuit board was used to connect the output of groups of four channels together, thereby increasing the maximum possible output to 6 mA per channel. Custom software in MATLAB was used to trigger and control stimulation.

Stimulation pulse trains consisted of charge-balanced, anodic-first square pulses, with symmetric anodic and cathodic phases. Stimulation was performed either in a monopolar configuration, with the ground electrode placed at a distant location such as on the skin at the shoulder or hip, or in a multipolar configuration with one or more local SCS contacts acting as the return path. Stimulation frequencies and pulse widths ranged 1–300 Hz and 50–1000 µs, respectively. The interphase interval was 60 µs.

### Recording perceptual responses

The methodology for recording perceptual responses, characterizing their psychophysical properties, and determining their stability at threshold have been detailed elsewhere^20^. Briefly, after a one-second stimulation train, subjects used a touchscreen interface^25^ developed in Python to document the location and perceptual quality of the evoked sensation. The location of the sensory percept was recorded using a free-hand drawing indicating the outline of the evoked percept on an image of the appropriate body segment (i.e., hand, arm or torso). The percept quality was recorded using several descriptors that have been used previously to characterize evoked percepts^26,27^.

### Motor control of prosthetic hand

Subjects performed an object discrimination task using a sensorized DEKA hand (Mobius Bionics) or a virtual representation of that hand, rendered in MuJoCo^28,29^. Because neither subject was a regular prosthesis user and time available for training was limited, each subject controlled the aperture of the prosthetic hand using a customized control signal. Subject 1 had a high trans-humeral amputation and did not have sufficient musculature in the residual limb to achieve reliable myoelectric control of the prosthesis. Instead, she wore a Data Glove (Fifth Dimension Technologies, 5DT) on her contralateral, intact hand, and the grasp aperture from the Data Glove was used to proportionally control the aperture of the real and virtual DEKA hand. Subject 2 had stroke-induced paralysis in her contralateral arm and could not use the Data Glove to control the DEKA hand. Because of this limitation and to more closely match the myoelectric approach that is commonly used clinically, bipolar surface EMG was recorded from the residual muscle in the ipsilateral forearm to control the prosthesis. EMG data were recorded at 2,000 Hz, high-pass filtered at 10 Hz, downsampled to 50 Hz by computing the moving average (20 ms bin size) and rectified. This rectified EMG signal was normalized to the peak EMG recorded during a maximum voluntary contraction prior to each session. Studies have shown that even in unilateral stroke, there is evidence of motor impairment in the ipsilateral limb; Subject 2 had difficulty achieving reliable prosthesis control, even with simple control schemes such as parallel dual-site control^30,31^. As such, we implemented a highly simplified control scheme, in which the subject was instructed to attempt a hand grasp, and when this signal crossed a manually defined threshold, the DEKA hand was commanded to close at a constant velocity of 15 degrees/s. Any time the processed EMG signal was below threshold, the hand was commanded to open at a constant velocity of 30 degrees/s. This behavior is similar to a normally-open body-powered prosthesis and a similar approach has been used previously by others to evaluate myoelectric robotic control^32,33^.

### Real-time somatosensory feedback via SCS

A subset of SCS electrodes that evoked focal percepts localized to the phantom hand and fingertips were used to provide real-time somatotopically-matched feedback during the object discrimination task. Sensors embedded in the fingers of the DEKA hand or virtual sensors in MuJoCo measured the force generated upon contact with the presented object. The maximum sensor force across the index, middle, and ring finger was mapped to an SCS electrode such that the receptive field of the evoked percept overlapped with the location of the sensors in the hand (Supplementary Figure 1). Custom software was written in MATLAB (Mathworks, Natick, MA) to process the grasp force and control stimulation in real-time with an update rate of 50 Hz.

Sensor signals were low-pass filtered using a 4^th^ order Butterworth filter with a cutoff at 4 Hz. We implemented both a linear and an exponential stimulation encoding scheme between grasp force and stimulus amplitude. Subject 1 performed the object discrimination task using both stimulation encoding schemes. Her performance using each encoding scheme is reported separately. Subject 2 performed the object discrimination task using the linear encoding scheme only. For the linear encoding scheme, the grasp force was first normalized to a scale ranging from 0 to 1 as shown in equation 1,

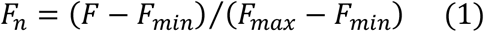

where, *F*_*n*_ is the normalized grasp force, *F* is the instantaneous grasp force, and *F*_*max*_and *F*_*min*_are the upper and lower limits of the grasp force measured by the sensor, respectively. The instantaneous stimulation amplitude (*A*) was determined as shown in equation

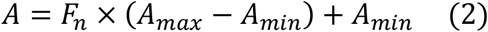

where, *A*_*max*_and *A*_*min*_are the upper and lower limits of the stimulus amplitude.

For the exponential encoding scheme, the instantaneous stimulation amplitude (*A*) was determined as shown in equation 3:

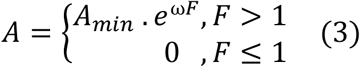

where, *F* is the instantaneous grasp force measured by the sensor, ω is an empirically assigned scaling factor (0.005 to 0.025), and *A*_*min*_is the lower limit of the stimulus amplitude.

### Object discrimination task design

The overall goal of this study was to provide functionally relevant somatosensory feedback. Due to the limited duration of these experiments, several parameters (such as the choice of stimulation encoding scheme, grasp force threshold, object geometry) were adjusted empirically across testing sessions and between subjects to improve the quality of the artificial somatosensory feedback. Table 1 provides a summary of the control scheme and number of object presentations for the physical and virtual object discrimination task for Subjects 1 and 2. Below, we provide more detail about the implementation and assessment of each of these tasks.

**Table 1:**
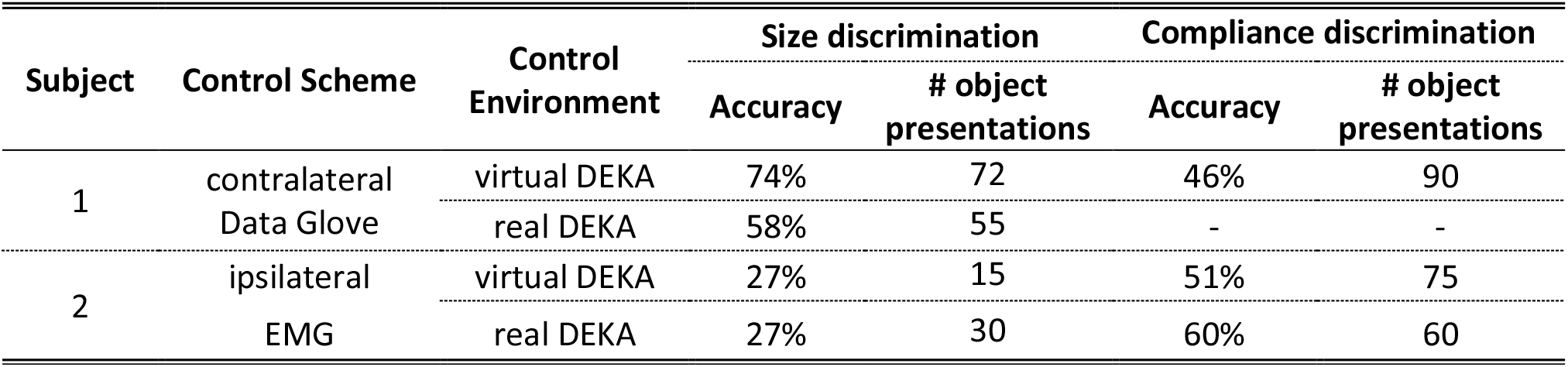
Summary of performance on object discrimination task for each subject.

### Virtual DEKA hand in MuJoCo

Both subjects used a virtual representation of the DEKA hand to perform the object discrimination task in a virtual environment designed using MuJoCo^34,35^. Subject 1 was presented with 9 spheres of three different sizes (small, medium, and large) and compliances (soft, medium, and hard) (Figure 1A). The subject had 10 seconds to interact with the object without visual feedback and an exponential stimulation encoding scheme between sensor force and stimulation was used for all object presentations. For the size discrimination task, each sphere was presented 8 times in random order resulting in a total of 72 object presentations. Both size and compliance were varied across trials, however, the subject was only asked to identify object size. Each size was presented 24 times. For the compliance discrimination task, each object was presented 10 times in random order resulting in a total of 90 object presentations.

Both size and compliance were varied across trials, however, the subject was only asked to identify object compliance. Each compliance level was presented 30 times.

**Figure 1:**
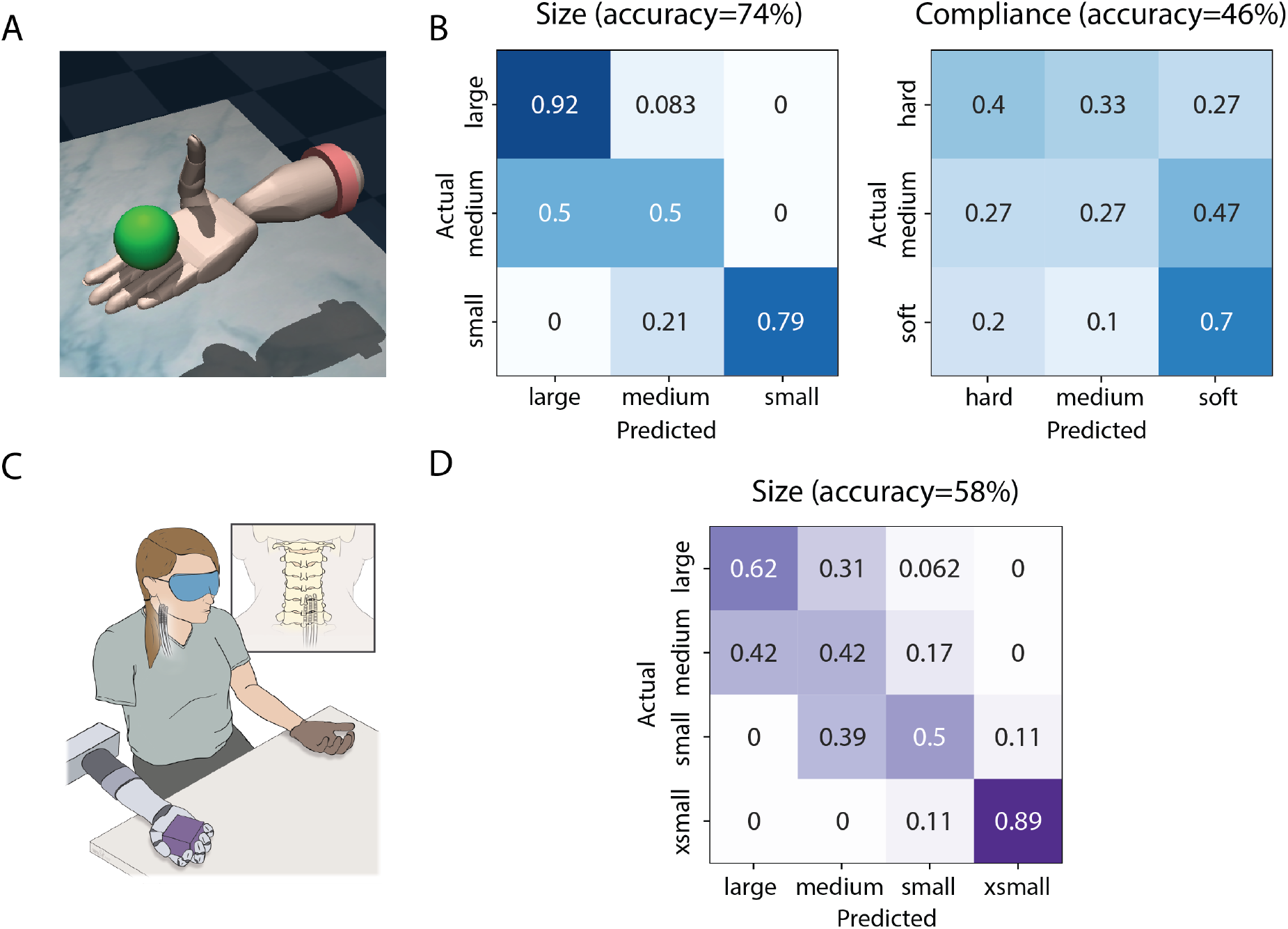
Object discrimination results for Subject 1. This subject used the DataGlove to control both virtual and physical prosthetic hands. A) Representation of the DEKA hand in the MuJoCo virtual environment with a spherical object. B) Confusion matrices for the object discrimination task using the virtual DEKA hand and an exponential stimulation encoding scheme. C) Experimental setup for the object discrimination task with the physical DEKA hand and DataGlove. D) Confusion matrix for the object size discrimination task using a linear stimulation encoding scheme. The compliance discrimination task with the DEKA hand was not performed for this subject.

For Subject 2, cylinders of three different sizes and compliance levels were presented (Figure 2A). The subject had 10 seconds to explore the object without visual or auditory feedback. A linear encoding scheme between sensor force and stimulation was used for all object presentations. For the compliance discrimination task, three different compliances (all with large size) were presented 25 times in random order resulting in a total of 75 object presentations.

**Figure 2:**
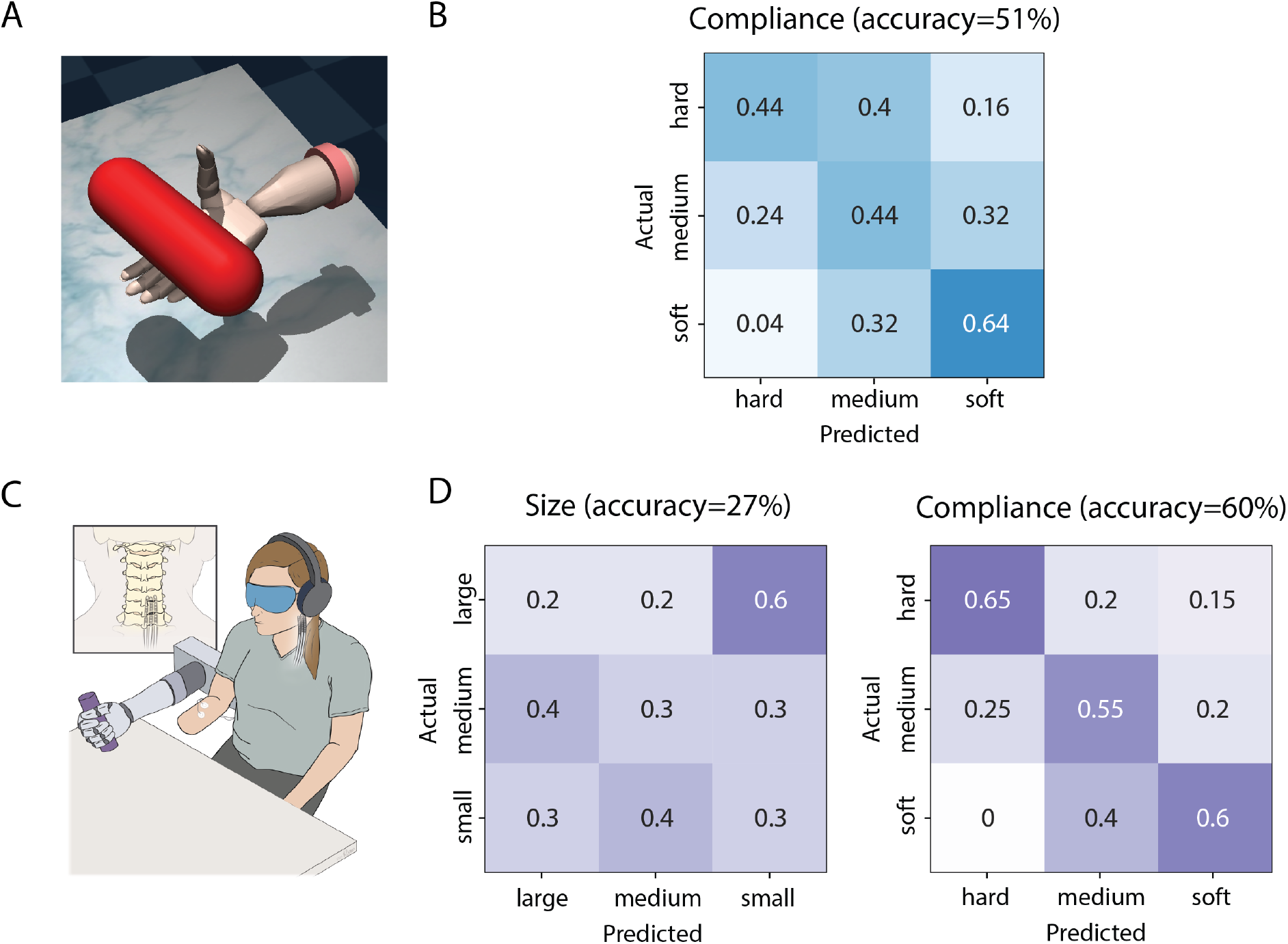
Object discrimination results for Subject 2. This subject used ipsilateral EMG signals to control closing of both the virtual and physical prosthetic hands. A) Representation of the DEKA hand in the MuJoCo virtual environment with a cylindrical object. B) Confusion matrix for the compliance discrimination task performance with the virtual DEKA hand using a linear stimulation encoding scheme. The size discrimination task with the virtual DEKA hand was not performed for this subject. C) Experimental setup for the object discrimination task with the physical DEKA hand and ipsilateral EMG electrodes. D) Confusion matrices for the object discrimination task using a linear stimulation encoding scheme.

### Physical DEKA hand

For the physical DEKA hand, Subject 1 was presented with cubes of four different sizes (extra small, small, medium, and large), all made from the same foam rubber material (Figure 1C). The order of presentation was randomized, and the subject performed the task without visual feedback. A timeout for object exploration was not enforced, however the subject never explored an object for more than 10 seconds. A linear encoding scheme between sensor force and stimulation was used for 9, 18, 12, and 16 presentations of the four sizes, respectively.

Subject 2 was presented with cylinders of three different sizes (small, medium, and large) or compliances (soft, medium, and hard) (Figure 2C). The design of these objects was changed from cubes to cylinders to reduce the slippage between the fingers and the corners of objects that we sometimes observed in Subject 1. The subject was given 20 seconds to explore each object.

For the size discrimination task, objects of three different sizes (all with hard compliance) were presented 10 times in random order resulting in a total of 30 object presentations. For the compliance discrimination task, three different compliances (all with medium size) were presented 20 times in random order resulting in a total of 60 object presentations. The subject performed the task without visual feedback while wearing noise-cancelling headphones. A linear encoding scheme between sensor force and stimulation was used for all object presentations.

### Statistical Analysis

To explore which features of the task were most correlated with successfully identifying object size or stiffness, we characterized the stimulation onset delay, peak stimulation amplitude, and the rate of change of stimulation for each object presentation. Additionally, for Subject 1 we characterized the contralateral hand aperture for each object presentation. For each subject, separate multivariate analyses of variance (MANOVA) were performed for object discrimination tasks involving the real and virtual DEKA hand. Size and compliance were the independent variables and hand aperture at stimulation onset, peak amplitude of stimulation and the rate of change of stimulation were the dependent variables. Subsequent univariate analysis (ANOVA) was carried out for each independent variable and *post-hoc* (Tukey-HSD) tests were carried out for dependent variables that displayed a significant effect. The Pearson correlation coefficient (*p*) between dependent variables that showed a significant difference across all objects was used to plot standard deviational ellipses. This allowed us to identify how object size and compliance were encoded through stimulation and postulate on the strategy employed by each subject during the object discrimination task. All statistical analyses were carried out using the *statsmodel* package in Python^36^.

## Results

### SCS evokes sensory percepts localized to the missing limb

We selected a subset of electrodes that evoked percepts in the phantom hand to provide somatotopically-matched sensory feedback in real-time as subjects interacted with objects of varying size and compliance. Supplementary Figure 1C shows representative sensory percepts evoked by these electrodes that were localized to the missing hand in each subject. A complete overview of the quality, stability, and psychophysical properties of percepts evoked via SCS has been detailed previously^20^.

### SCS provides functionally relevant somatosensory feedback

#### Subject : Object discrimination task performance

In the MuJoCo virtual environment (Figure 1A), Subject 1 was most successful in determining the size of the objects, with an overall accuracy of 74% (Figure 1B) across all 72 object presentations. The highest overall accuracy within a single set of 5 random presentations of each object size was 94%. Across multiple sessions, the subject correctly identified large and small objects (92% and 79% accuracy respectively) more often than medium objects (50%; chance level = 33%). Interestingly, when the subject misidentified medium-sized objects, they were always incorrectly identified as large-sized objects. When performance on the size discrimination task was analyzed for each object compliance (Supplementary Figure 2A), Subject 1 had the highest accuracy when presented with soft objects (95%) as opposed to medium and hard objects (71 % and 54%). Objects with stiffer compliance were frequently misidentified as larger sizes.

The subject was less accurate when determining object compliance with an overall accuracy of 46% (chance level = 33%), across all 90 object presentations. The peak performance within a single set of 6 random presentations of each object compliance was 50%. However, the subject was consistently more successful in identifying the soft object (70% accuracy, 36% false positive rate). Furthermore, when identifying object compliance for different object sizes (Supplementary Figure 2B), the subject performed below chance levels when presented with small objects (overall accuracy 13%) but performance improved for large objects (67% accuracy).

Interestingly, for medium and large objects the subject correctly identified soft objects with a high accuracy (100% and 80%) but she could only correctly identify all three compliances at above chance levels for the large object (70%, 50%, and 80% for soft, medium, and hard respectively).

The subject also used a physical DEKA hand to explore and identify objects of four different sizes with an overall accuracy of 58% (Figure 1C) across all 55 object presentations. The subject achieved accuracy rates of 62% and 89% with the largest and smallest objects (chance level = 25%), respectively. The confusion matrix (Figure 1D) shows that, for the two intermediate sizes, the subject commonly misidentified them as objects of adjacent larger sizes. This trend was similar to the misidentification of intermediate-sized objects observed in the virtual environment.

#### Subject 2: Object discrimination task performance

Subject 2 used the virtual DEKA hand to identify object compliance with an overall accuracy of 51% (Figure 2B). The highest overall accuracy within a single set of 5 random presentations of each object size was 60%. However, objects of adjacent compliance were frequently misidentified and only the soft objects were identified with an accuracy greater than 50%. Subject 2 also used the physical DEKA hand to identify object compliance with an overall accuracy of 60%. The confusion matrix in Figure 2D shows that for this task, the subject was able to identify the soft, medium, and hard objects with accuracies of 60%, 55%, and 65%, respectively. Performance on the compliance discrimination task was similar for small (67%), medium (60%) and large (54%) sized objects (Supplementary Figure 3).

Compared to compliance discrimination, the subject identified object size with a lower overall accuracy (27%, approximately at chance level) across 30 object presentations. The false positive rates for small, medium, and large objects were 20%, 40% and 50% respectively.

### Object size and compliance are encoded by independent stimulation features

We performed a post-hoc analysis of the sensor signals recorded during the object discrimination task to determine which features were most strongly correlated with subjects’ ability to accurately identify object compliance or size.

#### Subject 1

For Subject 1, when using the virtual DEKA hand, the contralateral hand aperture at stimulation onset was significantly different (p<0.01) for each object size. There was no overlap in the standard deviational ellipses for grasp aperture for all object presentations (Figure 3A).

**Figure 3:**
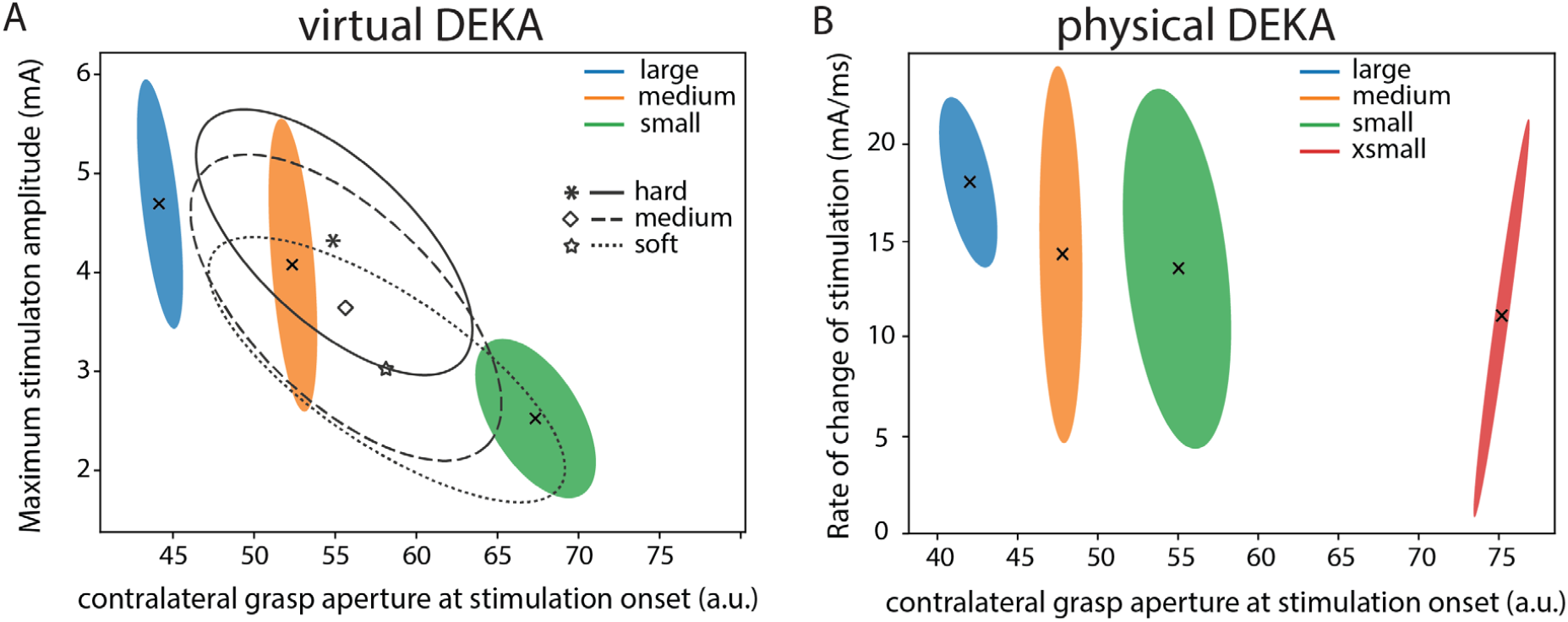
Salient features of stimulation correlate with subjects’ ability to discriminate object size or stifness. Standard deviational ellipses for A) the maximum stimulation amplitude and contralateral (DataGlove) grasp aperture at stimulation onset when using the virtual DEKA hand and B) the rate of change of stimulation and contralateral grasp aperture at stimulation onset, when using the physical DEKA hand for Subject 1.

Subject 1 may have attended to the aperture of her intact contralateral hand at stimulation onset to reliably perform the size discrimination task. It is also possible that Subject 1 utilized the timing of object contact to determine object size. Analysis of the lag between the onset of the grasp command and the onset of stimulation revealed that the delay in stimulation onset was significantly different when comparing small objects to either large or medium objects only (Supplementary Figure 4). Therefore, the timing of stimulation onset and contralateral grasp aperture both may have contributed to her performance on this task. The combination of multiple features may also explain the subject’s high overall accuracy on the size discrimination task (74% accuracy).

Further univariate analysis of object compliance revealed that the rate of change of stimulation was significantly different (p<0.01) when comparing hard objects to either soft or medium objects while the peak stimulation amplitude was significantly different for hard and soft objects only (p<0.01) (Supplementary Figure 4). Subject 1 may have attended to these features of stimulation to identify hard and soft objects (70% and 40% respectively) with a greater accuracy than objects with medium compliance (27%).

With the physical DEKA hand for Subject 1, object size was the only independent variable since all objects had a medium compliance. Similar to observations with the virtual DEKA hand, the contralateral hand aperture at stimulation onset was significantly different (p<0.01) for each object size and there was no overlap in the distribution of contralateral grasp apertures across all object presentations (Figure 3B). Further analysis of the lag between the onset of the grasp command and the onset of stimulation revealed that the delay in stimulation onset was significantly different when comparing extra small objects to either large or medium objects only (Supplementary Figure 4). This result indicates that Subject 1 could identify object size reliably from contralateral hand aperture at stimulation onset alone.

#### Subject 2

For Subject 2, when using the virtual DEKA hand, object compliance was the only independent variable since all objects were of medium size. A one-way ANOVA and subsequent post-hoc analysis confirmed that there was a significant difference in the rate of change of stimulation for each compliance (p<0.01). For all object presentations there was no overlap in the standard deviational ellipses for the rate of change of stimulation (Figure 4A). This result indicates that the subject may have relied on the rate of change of stimulation to determine object compliance. However, the overall low accuracy on the compliance detection task (51%) may be attributed to inherent difficulty in reliably detecting this stimulation feature.

**Figure 4:**
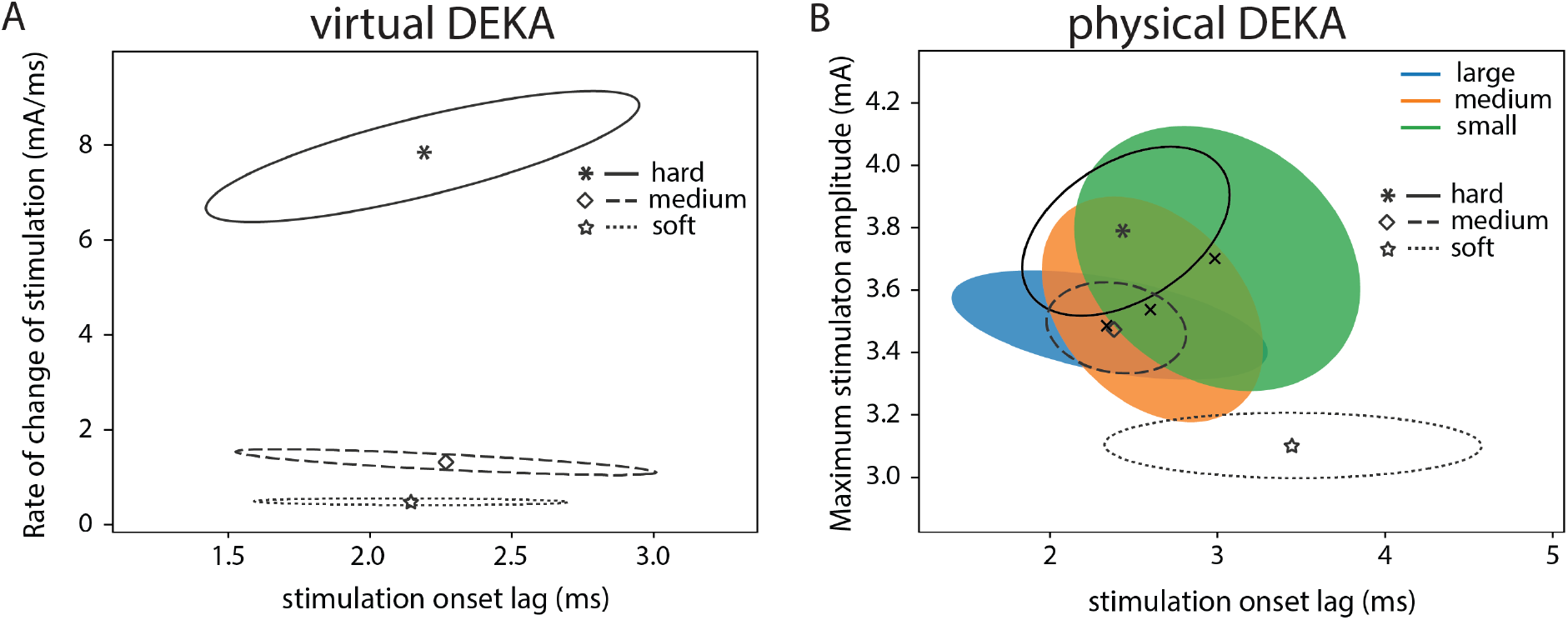
Salient features of stimulation correlate with subjects’ ability to discriminate object size or stifness. Standard deviational ellipses for A) the rate of change of stimulation and stimulation onset lag when using the virtual DEKA hand and B) the maximum stimulation amplitude and stimulation onset lag, when using the physical DEKA hand for Subject 2. The color of the ellipses represent object size and the line style represents object compliance. The centroid of each standard deviational ellipse represents the mean of the distribution for each object size and compliance.

For the object discrimination task with the physical DEKA hand, the rate of change of grasp force was similar for all objects since the hand was commanded to close at a constant velocity. Similar to observations in Subject 1, the aperture of the DEKA hand at stimulation onset was significantly different (p<0.01) for each object size (Supplementary Figure 5).

However, there was no statistically significant relationship between grasp aperture and any feature of stimulation. For all object presentations, there was significant overlap in the standard deviational ellipses for the lag between the onset of the grasp command and the onset of stimulation (Figure 4B). Object size could not be conveyed to the subject via this control scheme, which likely explains the subject’s decreased performance on the size discrimination task. However, univariate analysis showed that there was a statistically significant relationship between object compliance and the peak stimulation amplitude (p<0.01). Post-hoc tests showed that the peak stimulation amplitude was significantly different for all object compliances (p<0.01). Therefore, Subject 2 may have used stimulation amplitude to identify different object compliances.

## Discussion

### Subjects can use somatosensory feedback via SCS during an object discrimination task

In this study, we demonstrate that somatotopically-matched real-time feedback provided by SCS can be used by subjects to determine object size or compliance. Subject 1 was consistently more adept at determining object size (up to 74% accuracy) while Subject 2 achieved a higher accuracy level in determining object compliance (up to 60% accuracy). Both subjects could readily use the sensory feedback with minimal training. However, performance varied based on the environment (real or virtual DEKA hand) and the control strategy (contralateral grasp aperture control or ipsilateral linear velocity myoelectric control).

### Utility of somatosensory feedback is dependent on task design

While both subjects were able to incorporate sensory feedback into their use of a prosthetic device, success rates for identifying object size and stiffness were starkly different between subjects. Subject 1 had proportional control of the DEKA hand through a DataGlove on her contralateral intact hand. This control strategy provided her with information about grasp aperture through the intact proprioceptive pathways in the contralateral limb. Combined with the timing of stimulation onset, this proprioceptive information provided a reliable estimate of object size. Additionally, Subject 1 did not wear noise-cancelling headphones when performing the task using the real DEKA hand. However, performance on the object discrimination task was comparable using the virtual and physical DEKA hand, suggesting the presence or lack of audio feedback did not affect the experiment.

Conversely, Subject 2 used an EMG signal to control the closing velocity of the prosthetic hand such that grasping the object required a sustained finger/wrist flexion. In the absence of proprioceptive feedback, Subject 2 may have attended to the time delay between the onset of wrist/finger flexion and the onset of stimulation to infer the grasp aperture at object contact. However, the subject had no feedback about when the signal exceeded the threshold, leading to variability between the onset of flexion and the onset of grasp. This variability resulted in objects of different sizes having overlapping stimulation onset lags and likely explains her decreased accuracy on the size discrimination task.

Other studies have also shown that artificial sensation that conveys grasp aperture information is key to determining object size when using a prosthesis^12,15,37^. However, providing true proprioceptive information through artificial somatosensory feedback has consistently been a difficult challenge for somatosensory neuroprostheses. Most studies have provided grasp aperture information by remapping it to a tactile sensation^15^ or to a sensation of movement of a specific finger or joint^16^. It is plausible that a similar strategy employed with Subject 2 may have provided useful proprioceptive feedback to perform the size discrimination task accurately.

The object discrimination task for this study was intentionally designed to be flexible and did not constrain the subjects’ behavior to a rigid protocol. This resulted in multiple confounding factors that must be considered when attempting to explain overall task performance. Our results indicated that for both subjects, the rate of change of stimulation and peak stimulation amplitude encoded object compliance. However, only Subject 2 seemed to utilize this information to determine object compliance (up to 60% accuracy). Subject 2’s performance may be attributed to a better ability than Subject 1 in discriminating the rate of change of stimulation (using the virtual DEKA) and the peak stimulus amplitude (using the real DEKA). However, the control strategy employed in either subject may have also affected the utility of the feedback they received. When interacting with objects, Subject 1 made frequent ballistic movements of the contralateral hand that resulted in rapid changes in grasp aperture. It is possible that subtle differences in stimulation dynamics across objects of different compliance could not be detected for such short durations of stimulation (or object contact). In contrast, for Subject 2 a fixed closing velocity ensured that the dynamics of stimulation were consistent for objects with the same compliance. It is possible that controlling the closing velocity for Subject 1 may have provided a reliable estimate of compliance and improved performance on the compliance discrimination task.

Overall, features of stimulation encode specific physical properties of objects used during the task. However, the design of the task and control strategy of the prosthetic determine whether these features can be reliably detected by the subject.

### Considerations for closed loop prosthesis design

In this study, we demonstrated that SCS provides somatosensory feedback that subjects can use to identify the size or compliance of objects. However, there are several shortcomings that should be addressed in future work. The percutaneous SCS system described here was implanted for up to 29 days in both subjects. Initial experiments focused on mapping evoked percepts and studying their psychophysics^20^. This information is vital to determining the electrodes and stimulation parameters to use during the discrimination tasks studied here. However, this also constrained the amount of experimental time to study these functional tasks. Neither subject had prior experience using a prosthesis, and these tasks were performed after only limited training. It is likely that over time, subjects could learn to attend to specific changes in the stimulation and improve their performance on the object discrimination tasks. Future work should focus on tracking subject performance across multiple days.

Previously reported psychophysics data showed that both subjects could discriminate three specific intensity levels (82% and 79% accuracy) based on trains of stimulation that had three discrete amplitudes^20^. In the present study, somatosensory feedback was modulated in real-time. Therefore, subjects had to attend to stimulation dynamics (rate of change, peak amplitude) instead of the instantaneous intensity of the evoked percept. However, our psychophysics testing did not quantify subjects’ ability to detect changes in stimulation dynamics. In fact, identifying the stimulation encoding scheme that provided the best discrimination of different objects was a major challenge. Future work should focus on characterizing the threshold and just-noticeable difference for dynamic properties of stimulation to identify the optimal stimulation encoding scheme.

Additionally, it is worth noting that the object discrimination task used in this study and several other studies^13,15,16,38^ is essentially a modified magnitude discrimination task. When all other object properties are held constant, a single stimulation feature (e.g. rate of change of stimulation) can encode a distinct property of an object (e.g. deformation). Subjects that can perceive gradation in this feature will demonstrate higher accuracy. However, these results may not generalize to a real-world somatosensory neuroprosthesis. When presented with a novel object, the same features of stimulation may encode multiple physical properties (e.g. deformation and movement of an object) so more complex stimulation schemes may be required. Critically, studies that monitor subject performance during activities of daily living and novel interactions are necessary to characterize the functional utility of artificial somatosensory feedback^19,39,40^.

## Data Availability

All data produced in the present study are available upon reasonable request to the authors

## Acknowledgements

We would like to thank Howard Stein, Lauren Wilcox, Emily Bird, Bree Bigelow, and Debbie Harrington for their recruitment efforts, regulatory compliance, and clinical scheduling. We would also like to thank David Weir and Tyler Simpson for their engineering and technical support. Most importantly, we would like to thank the research subjects for their participation in this study. Research was sponsored by the U.S. Army Research Office and the Defense Advanced Research Projects Agency (DARPA) was accomplished under Cooperative Agreement Number W911NF-15-2-0016. The views and conclusions contained in this document are those of the authors and should not be interpreted as representing the official policies, either expressed or implied, of the Army Research Office or the U.S. Government. The U.S. Government is authorized to reproduce and distribute reprints for Government purposes notwithstanding any copyright notation hereon.

## Supplementary Figures

**Supplementary Figure 1:**
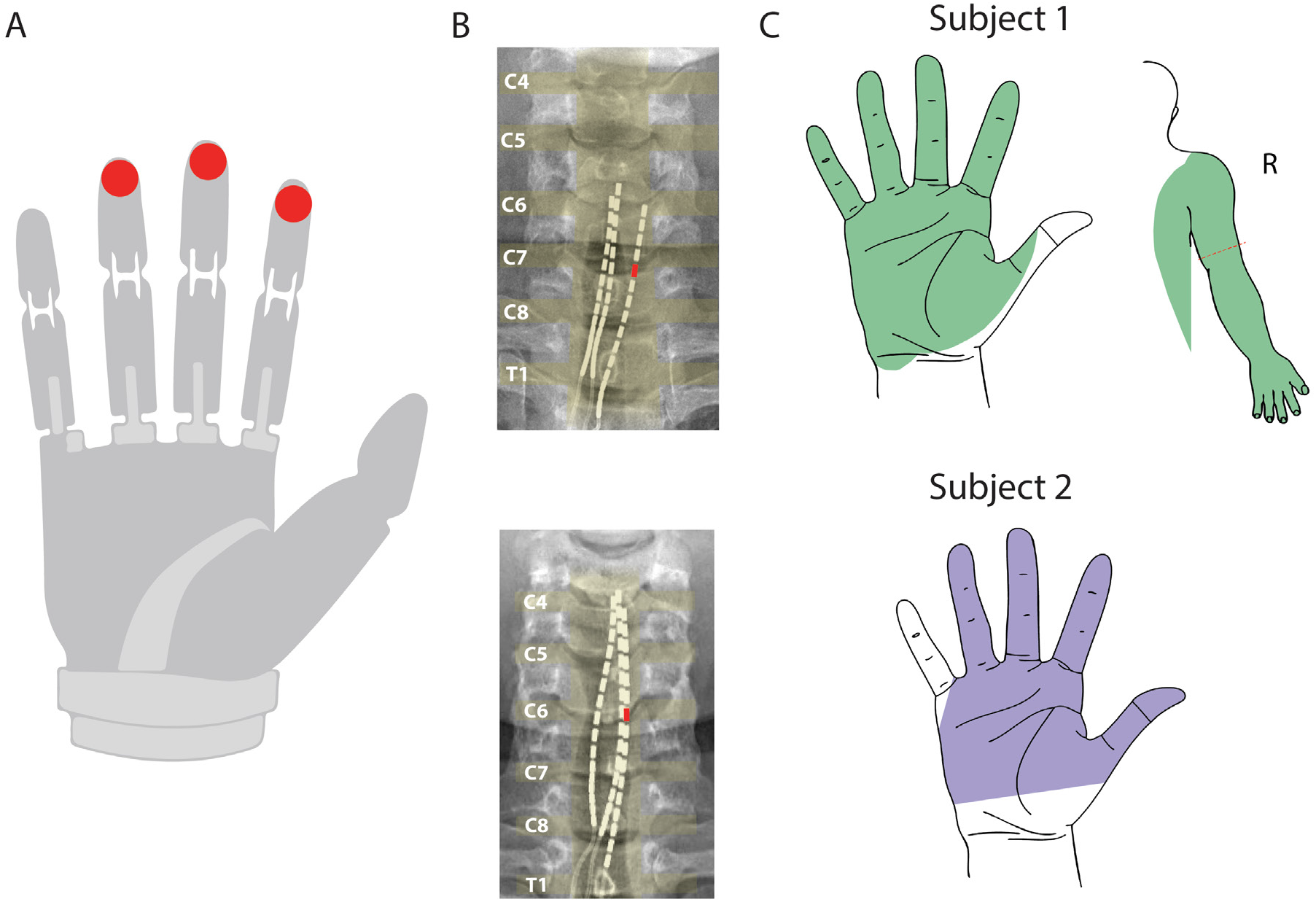
A) Schematic of grip force sensor location on the index, middle, and ring fingers of the DEKA hand. The maximum value across all three sensors was used to encode the stimulation amplitude. B) X-ray of lead position during closed loop testing sessions. The red rectangle denotes the stimulation electrode mapped to the grip sensors. C) Representative sensory percepts for Subjects 1 and 2 generated by electrodes from B. Colored areas represent the stable projected fields of the electrodes that were used during closed loop object discrimination tasks. Simultaneously evoked percepts in the residual arm are shown for Subject 1.

**Supplementary Figure 2:**
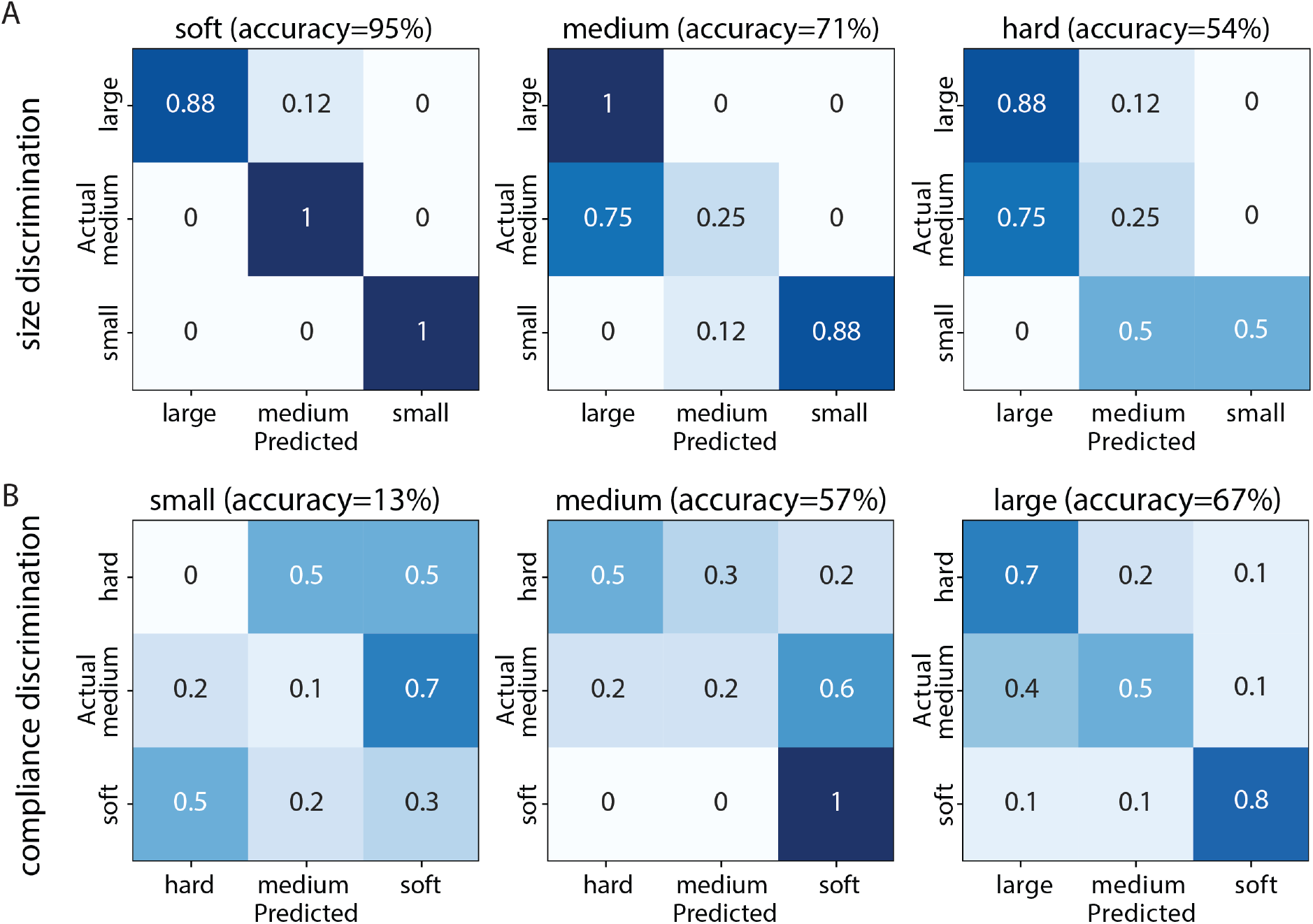
Confusion matrices for the performance of Subject 1 on the A) compliance discrimination task for each object size and the B) size discrimination task for each object compliance using the virtual DEKA hand.

**Supplementary Figure 3:**
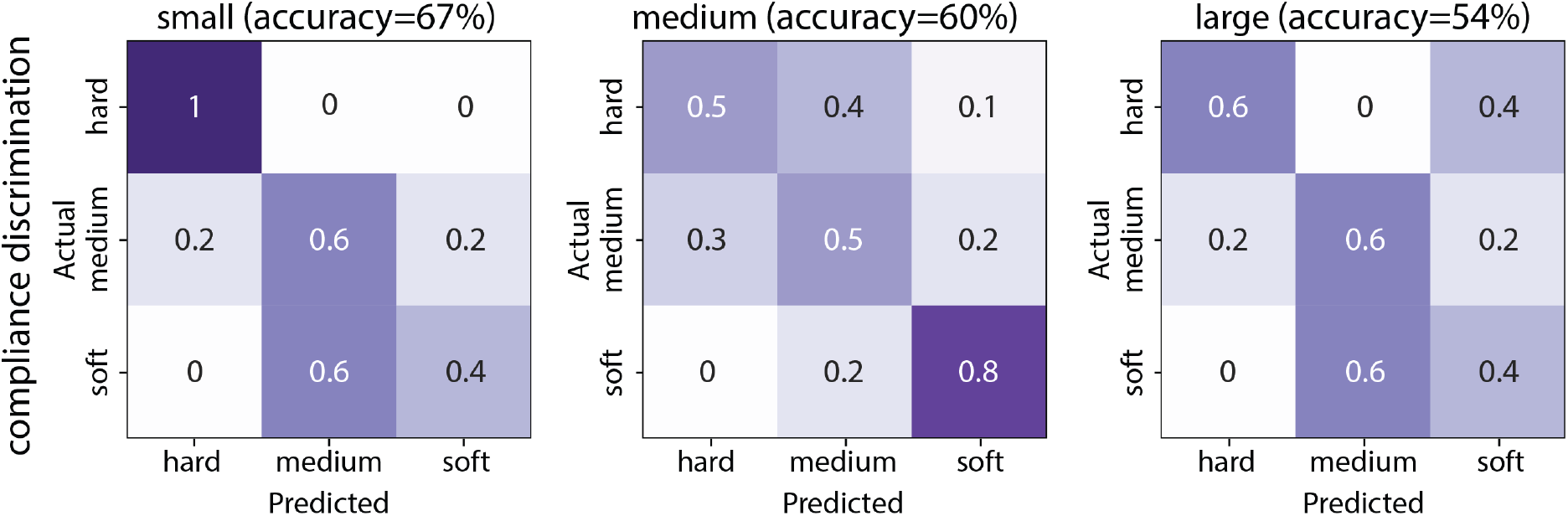
Confusion matrices for the performance of Subject 2 on the compliance discrimination task for each object size using the physical DEKA hand.

**Supplementary Figure 4:**
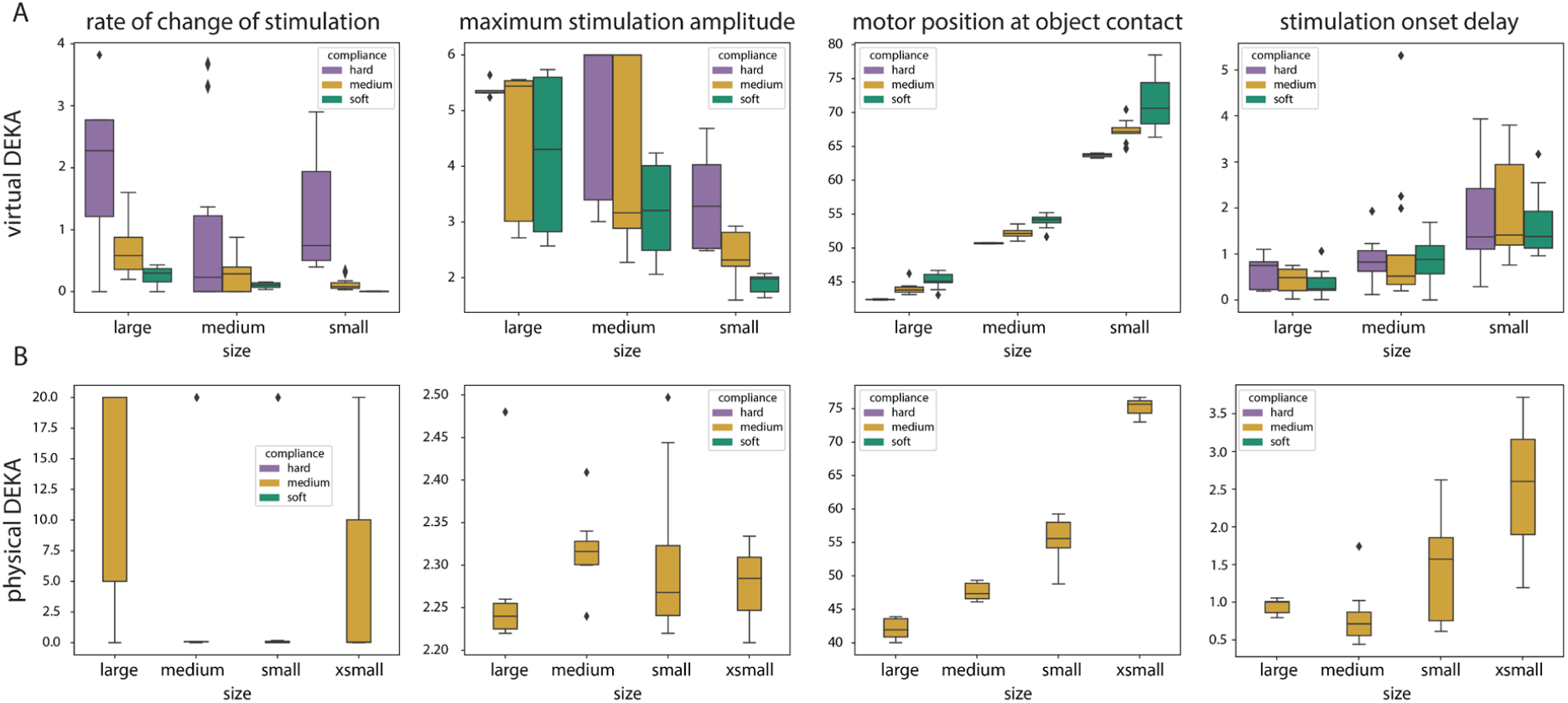
Distribution of rate of change of stimulation, peak stimulation amplitude, grasp aperture at stimulation onset and stimulation onset delay for all object presentations using the A) virtual and B) physical DEKA hand for Subject 1.

**Supplementary Figure 5:**
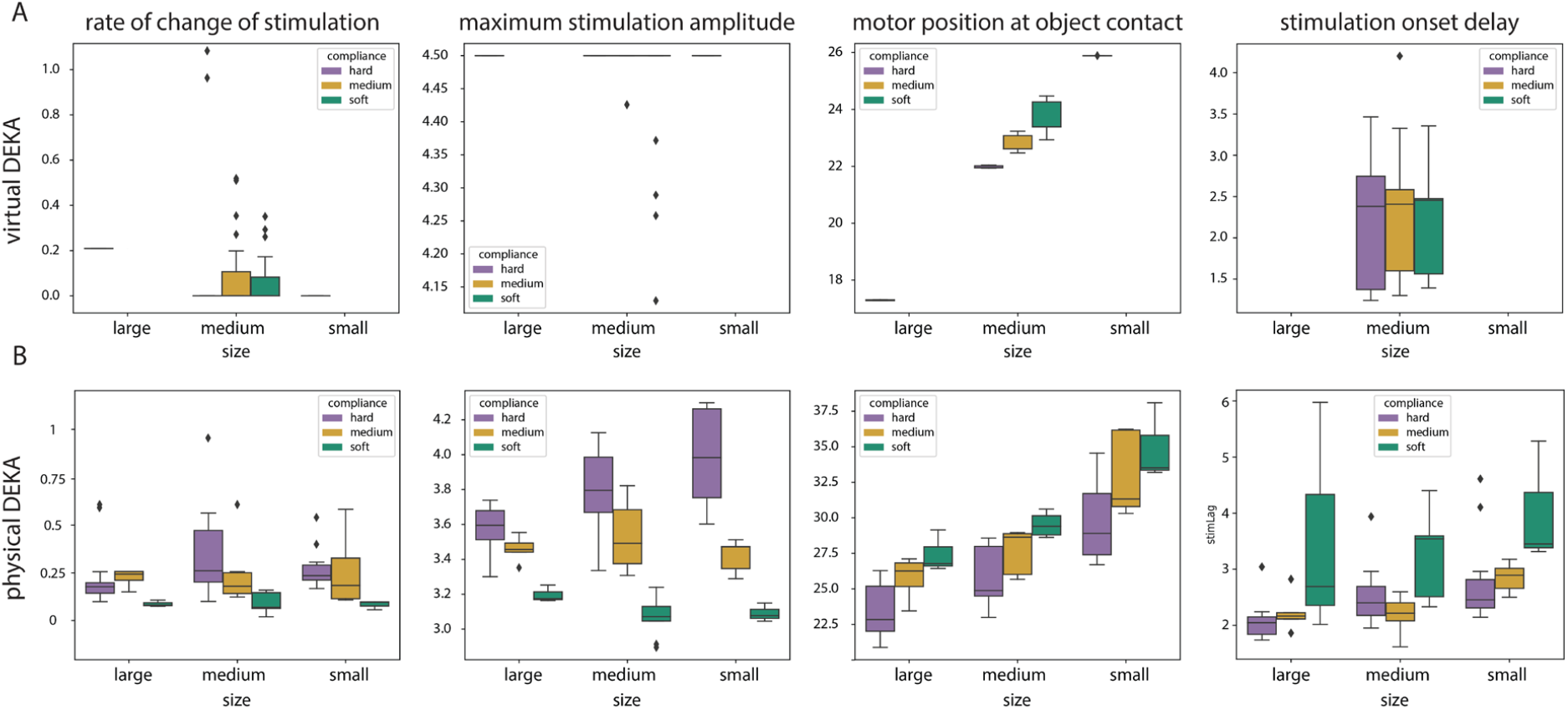
Distribution of rate of change of stimulation, peak stimulation amplitude, grasp aperture at stimulation onset and stimulation onset delay for all object presentations using the A) virtual and B) physical DEKA hand for Subject 2.

